# Early Feasibility of NIVA Score Decongestion Responsiveness: A Pilot Clinical and Preclinical Study

**DOI:** 10.64898/2026.07.13.26357910

**Authors:** Bret D. Alvis, Jeffrey Schmeckpeper, Aniket S. Rali, Jessica Huston, Stacy Tsai, Kaushik Amancherla, David Armstrong, Richa Gupta, Jonathan S. Whitfield, Rene Harder, Katharine Miller, Mackenzie Horne, Dawson Wervey, Romy Pein, Tara Isanaka, Marisa Case, Eric Wise, Bryce Perrien, Colleen Brophy, JoAnn Lindenfeld, Kyle Hocking

**Author notes:** **Corresponding author** Bret D. Alvis, MD, Vanderbilt University Medical Center, 422 K, Medical Arts Building, Nashville, TN 37232, USA.

## Abstract

Residual congestion is the principal driver of heart failure readmission, and reliable serial assessment of volume status remains an unmet clinical need. This study asked whether a wrist-worn, machine-learning–based device for non-invasive venous waveform analysis in heart failure (the NIVA_HF_ device), which produces an integer-scaled estimate of pulmonary capillary wedge pressure termed the NIVA Score, responds to acute changes in volume status.

Agreement between the NIVA Score and invasively measured pulmonary capillary wedge pressure at single time points has been established in a separate prospective, multi-site study; however, such static agreement does not establish whether the measure tracks dynamic decongestion. We therefore evaluated the directional responsiveness of the locked NIVA Score in two prespecified cohorts: hospitalized adults with acute decompensated heart failure undergoing routine intravenous diuresis, and a controlled porcine model of volume overload followed by diuresis. In eleven patients contributing thirteen paired measurements (mean net fluid balance −2.1 ± 1.0 L), NIVA Scores decreased significantly after diuresis (paired t-test, P = 0.04). In five pigs contributing twenty-four paired measurements, NIVA Scores decreased significantly after intravenous furosemide following crystalloid loading (P <0.01), and the direction of change was concordant with measured urine output in every animal. Statistical significance was reached in both cohorts despite modest sample sizes, indicating a measurable NIVA Score reduction with volume removal. In an exploratory analysis, the discharge NIVA Score yielded an area under the receiver-operating-characteristic curve of 0.85 (95% confidence interval 0.575–1.00; P = 0.04) for thirty-day readmission. Together, the significant, directionally concordant NIVA Score reductions across independent clinical and preclinical cohorts demonstrate that the device tracks acute decongestion and support its use for serial, non-invasive congestion monitoring; an adequately powered prospective study is the planned next step.

## INTRODUCTION

Residual congestion is the dominant driver of heart failure (HF) readmission, and serial assessment of volume status is central to the 2024 ACC Expert Consensus Decision Pathway on Clinical Assessment, Management, and Trajectory of Patients Hospitalized With Heart Failure [1]. Implanted hemodynamic monitors (CardioMEMS, Cordella) demonstrate that hemodynamically guided care reduces HF events, but invasive cost and procedural risk limit adoption [2,8]. Physical examination alone correlates poorly with invasively measured filling pressures [7], underscoring the need for an accurate, low-burden, repeatable measure of congestion that can be obtained serially outside the catheterization laboratory. A wrist-worn, machine-learning–based Non-Invasive Venous waveform Analysis for Heart Failure (NIVA_HF_) device estimates pulmonary capillary wedge pressure (PCWP) from peripheral venous waveforms via an integer-scaled output (the NIVA Score). Independent multi-site validation of NIVA Score agreement with invasively measured PCWP is reported separately [3], building on earlier observations that noninvasive venous waveform analysis correlates with intracardiac filling pressures [4,5].

Independent validation against a static reference (right heart catheterization) does not, by itself, establish whether a noninvasive congestion metric tracks dynamic changes in volume status, the property required for serial monitoring and for guiding decongestive therapy [6]. Here we report data on the directional responsiveness of the locked NIVA Score to acute decongestion in (Cohort 1) hospitalized adults with acute decompensated HF (ADHF) undergoing routine intravenous diuresis and (Cohort 2) a tightly controlled porcine volume-overload/diuresis model.

## MATERIALS AND METHODS

### Ethics statement

The clinical cohort was enrolled under an observational protocol approved by the University of Alabama at Birmingham Institutional Review Board, ceded through the Vanderbilt University Medical Center Institutional Review Board; all participants provided written informed consent prior to any study procedure. The porcine study was conducted under a protocol approved by the Vanderbilt University Medical Center Institutional Animal Care and Use Committee and in accordance with the ARRIVE guidelines for the reporting of animal research.

### Study design

The locked NIVA Score algorithm, independently validated against invasively measured PCWP in a separate prospective, multi-site study [3], was applied to two prespecified cohorts. No further algorithm training, tuning, or thresholding was performed for this study. Statistical significance was prespecified at P < 0.05, and all paired analyses were restricted to subjects or animals with both pre- and post-diuresis recordings. Analyses were performed in JMP Pro 18 (SAS Institute, Cary, NC, USA). Although no formal a priori sample-size calculation was performed, the prespecified paired comparisons of NIVA Score before and after decongestion reached statistical significance in both cohorts, indicating that the achieved sample sizes were sufficient to detect a within-subject response; the 30-day readmission analysis was exploratory.

### Cohort 1—ADHF pilot

Adult patients hospitalized with a clinical diagnosis of ADHF from 5/21/2025 to 10/30/2025 and receiving intravenous loop diuretics per routine clinical care were enrolled at a single academic medical center. After written informed consent, the wrist-worn NIVA_HF_ device was applied to the volar aspect of the wrist opposite any peripheral intravenous catheter [3 - 5]. Venous waveform recordings were obtained at two predefined time points: pre-diuresis (within 30 minutes of the diuretic dose) and post-diuresis (4 hours after the diuretic dose). Subjects were required to demonstrate ≥500 mL of net negative fluid balance over the interval. Treating clinicians were blinded to NIVA Scores, and no modifications were made to clinical management. Acquisitions were uploaded to a secure REDCap database and processed by the locked NIVA algorithm. Pre-versus post-diuresis NIVA Scores were compared by paired t-test. As an exploratory analysis, ROC discrimination of the discharge (post-final-diuresis) NIVA Score for 30-day HF readmission was evaluated.

### Cohort 2—Porcine volume-overload model

Five female Yorkshire pigs (~40 kg, 11–13 weeks) were studied. Animals were anesthetized (ketamine/xylazine/telazol induction, isoflurane maintenance), intubated, and instrumented with bladder, femoral arterial, and femoral venous lines. Bilateral NIVA_HF_ devices were placed on both upper extremities (**Figure 1**). Baseline NIVA Scores were recorded; animals then received 2,500 mL of warmed PlasmaLyte over ~25 minutes followed by a 10-minute acclimation period, and post-loading NIVA Scores were recorded. Animals subsequently received 200 mg intravenous furosemide and were monitored for 75 minutes with urine output recorded; final NIVA Scores were then obtained. Paired averaged peak-overload versus post-diuresis NIVA Scores were compared by paired t-test.

**Figure 1.**
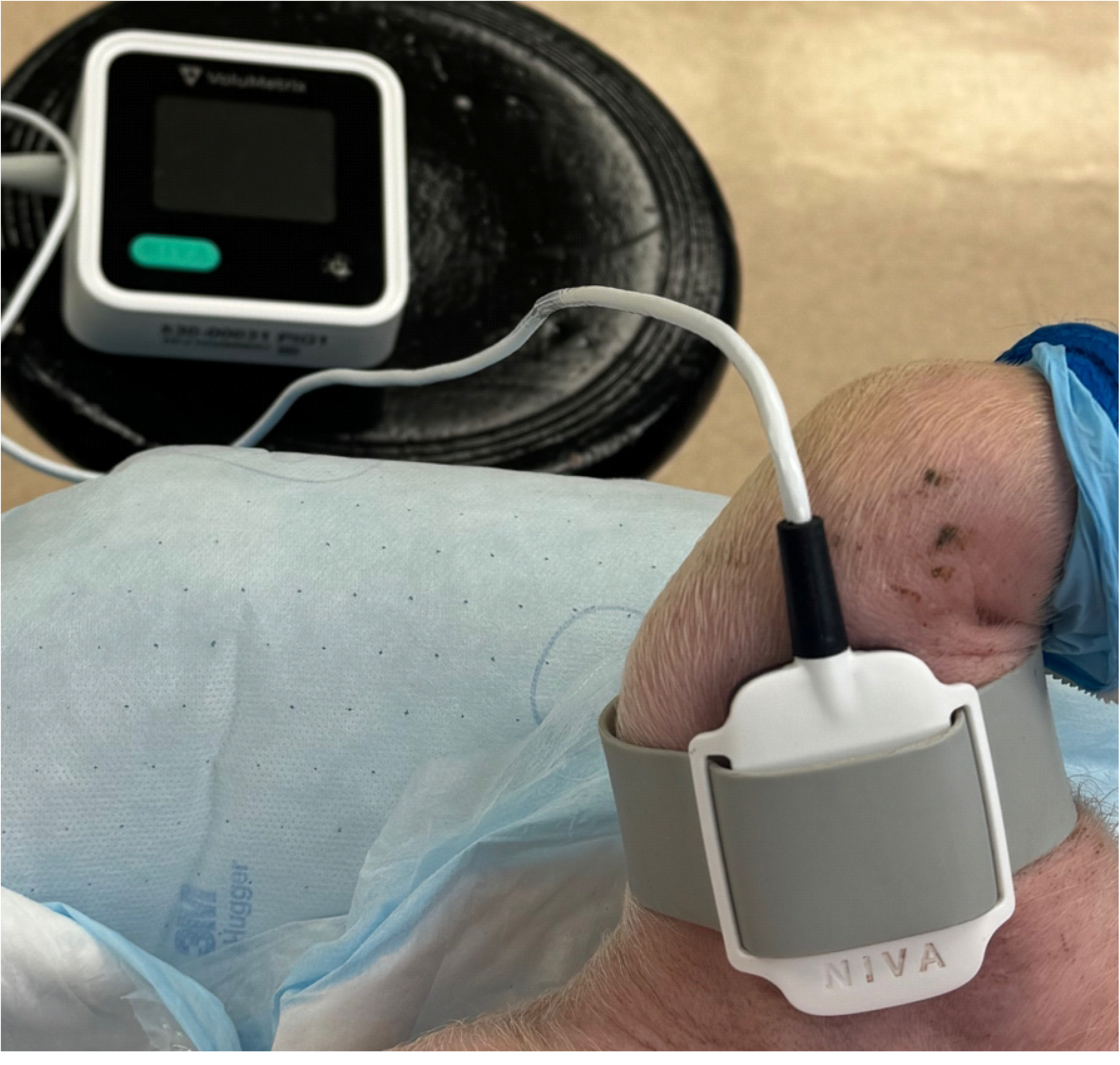
The NIVA_HF_ device. Bilateral NIVA_HF_ devices on the upper extremities of an anesthetized female Yorkshire pig in the volume-overload/diuresis model, Cohort 2. This same technology (NIVA_HF_ base unit and wristband) but different physical device were used in Cohort 1—heart failure patients undergoing diuresis. Abbreviations: NIVA_HF_, Non-Invasive Venous Waveform Analysis for Heart Failure; NIVA, Non-Invasive Venous Waveform Analysis.

## RESULTS

### Cohort 1

Eleven hospitalized ADHF patients contributed 13 paired pre- and post-diuresis NIVA measurements. Mean net fluid balance over the measurement interval was −2.1 ± 1.0 L. NIVA Scores decreased significantly following diuresis (paired t-test, P = 0.04; **Figure 2A**); this statistically significant within-patient reduction demonstrates a NIVA Score response to acute decongestion, and the achieved sample size was sufficient to detect it. In an exploratory analysis, the discharge (post-final-diuresis) NIVA Score showed an area under the ROC curve of 0.85 (95% CI 0.575–1.00; P = 0.04) for 30-day HF readmission (**Figure 2B**). At a discharge NIVA Score threshold of ≥20, the area under the curve for 30-day readmission was 0.94 (95% CI 0.813–1.00; P = 0.01), with sensitivity 60% and specificity 100% (positive predictive value 100%; negative predictive value 61%). Because this readmission cohort was small and the associated confidence intervals wide, the prognostic analysis is considered preliminary.

**Figure 2.**
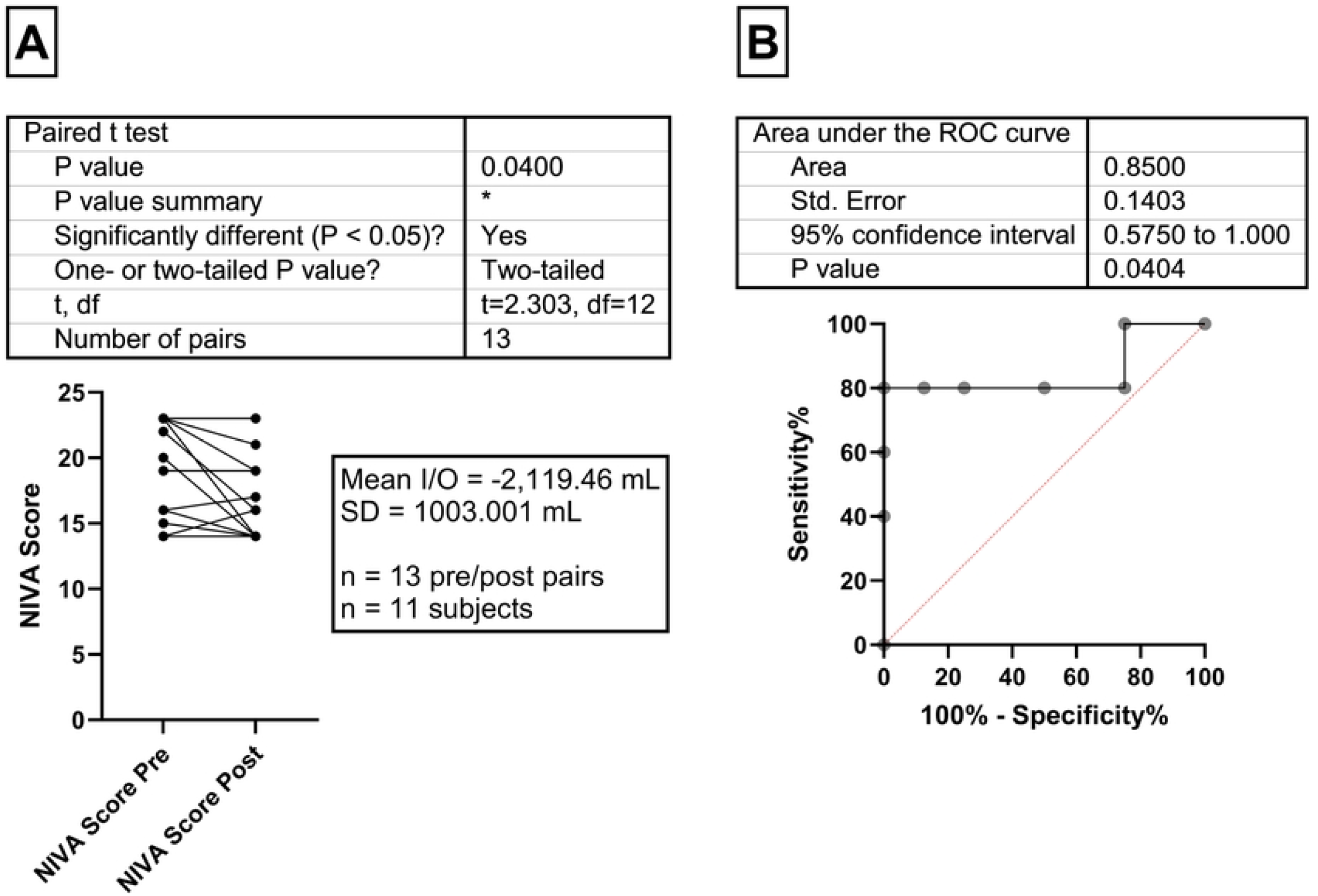
NIVA Score response to clinical diuresis in hospitalized ADHF patients. (A) Paired pre-versus post-diuresis NIVA Scores (n = 13 paired measurements in 11 patients) decreased significantly following routine intravenous loop diuretic therapy (paired t-test, P = 0.04). (B) Exploratory ROC analysis of the discharge (post-final-diuresis) NIVA Score for 30-day HF readmission (n = 13) yielded an area under the curve of 0.85 (95% CI 0.575–1.00; P = 0.04); this exploratory prognostic analysis should be interpreted with caution given the small cohort and wide confidence interval. Abbreviations: NIVA, Non-Invasive Venous Waveform Analysis; ADHF, Acute Decompensated Heart Failure; ROC, Receiver Operating Characteristic; AUC, Area Under the Curve; HF, heart failure; CI, confidence interval.

### Cohort 2

Five pigs contributed 24 paired NIVA Score measurements from bilateral wrist devices. Following volume loading (2.5 L) and subsequent diuresis (200 mg furosemide; mean urine output 1.5 ± 0.3 L), NIVA Scores decreased significantly (P <0.0007; **Figure 3**). Directional concordance between urine output and NIVA Score reduction was observed in all five animals.

**Figure 3.**
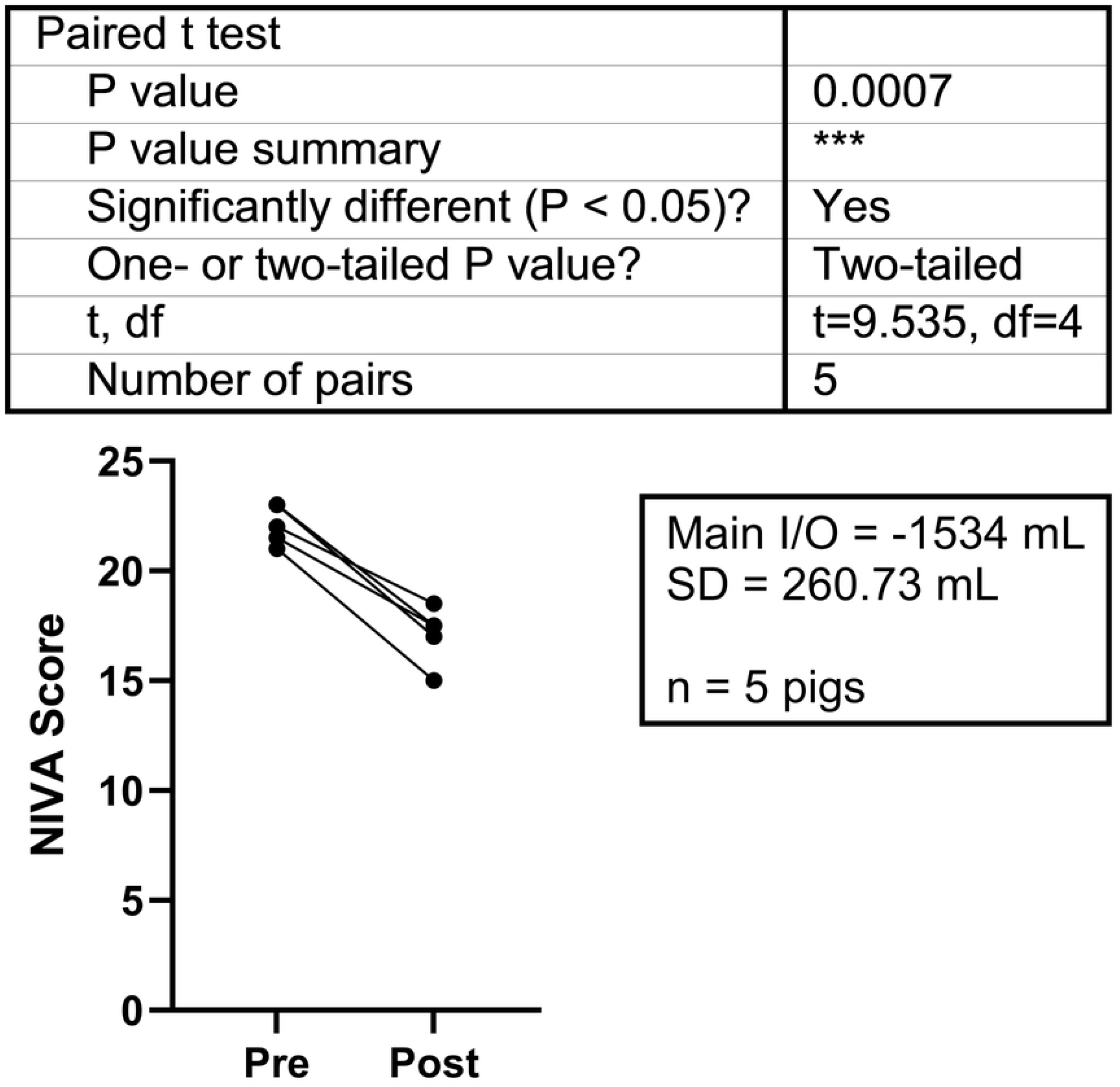
NIVA Score response to controlled volume loading and acute diuresis in a porcine model. Paired averaged (right and left upper extremity) NIVA Scores obtained at peak volume overload (after 2.5 L isotonic crystalloid loading) versus after 200 mg intravenous furosemide demonstrated a significant decrease (P <0.01; n = 5 animals, 24 paired measurements). Mean urine output during the diuretic phase was 1.5 ± 0.3 L. Directional concordance between urine output and NIVA Score reduction was observed in all five animals. Abbreviations: NIVA, Non-Invasive Venous Waveform Analysis; SD, standard deviation.

## DISCUSSION

In two prespecified cohorts (one clinical, one preclinical) the locked NIVA Score decreased significantly after acute decongestion, with statistical significance reached in both despite modest sample sizes. The porcine model provides mechanistic confirmation under tightly controlled preload manipulation, whereas the ADHF pilot demonstrates a directionally consistent signal under heterogeneous, real-world clinical conditions. Together these findings support the concept that the NIVA Score reflects dynamic changes in volume status rather than a fixed physiologic phenotype, complementing the static agreement with invasively measured PCWP reported separately in a prospective, multi-site study [3].

The convergence of these two datasets is mechanistically informative. Peripheral venous waveforms encode information about central venous return and cardiac filling that is transmitted through the venous system, and the NIVA Score is derived from features of that waveform that covary with intracardiac filling pressures [4,5]. In the porcine model, preload was manipulated across a wide range under anesthesia with continuous instrumentation, isolating the effect of acute volume change from the confounders present at the bedside; the observation that every animal exhibited a fall in NIVA Score concordant with measured urine output argues that the signal tracks intravascular volume rather than an incidental correlate of the loading protocol. In the ADHF cohort, the same directional response emerged despite heterogeneous diuretic regimens, comorbidities, and body habitus, suggesting that the effect is robust to the variability of real-world care. That a metric validated cross-sectionally against static PCWP also moves appropriately with acute decongestion is the central requirement for any tool intended to guide therapy over time, and it distinguishes a responsive biomarker from one that merely classifies a fixed hemodynamic state.

These observations position the NIVA_HF_ device within an evolving landscape of congestion assessment. Implantable pulmonary-artery and left-atrial pressure monitors reduce heart-failure events by enabling pressure-guided titration, but their cost, procedural risk, and need for implantation restrict their use to selected patients [2,8]. Physical examination and natriuretic peptides remain imperfect surrogates for filling pressure, and clinical assessment is frequently discordant with invasive hemodynamics [7]. A wrist-worn, non-invasive measurement that can be repeated as often as needed, at the bedside during hospitalization and potentially in the ambulatory setting, could extend the benefits of hemodynamically informed care to a broader population without an implanted device. The association between a higher discharge NIVA Score and 30-day readmission, though derived from a small cohort, is directionally consistent with this framing and supports the hypothesis that residual congestion detectable non-invasively at discharge identifies patients at elevated near-term risk. Whether serial NIVA-guided decongestion improves outcomes, and what thresholds are clinically actionable, are the central questions for future prospective work.

Several limitations apply. Both cohorts were modest in size and were not accompanied by a formal a priori power calculation; nonetheless, the prespecified paired comparisons of NIVA Score before and after decongestion reached statistical significance in both the clinical and preclinical cohorts, indicating that these sample sizes were sufficient to demonstrate a genuine within-subject response rather than a chance trend. Residual physiologic variability during clinical diuresis, arising from redistribution of third-space fluid, heterogeneity of cardiac function, and comorbidities, may attenuate or add noise to the effect in larger, more diverse populations, and the magnitude of the response therefore warrants confirmation at scale. The 30-day readmission analysis is the most preliminary element, deriving from few events with a wide confidence interval, and should be regarded as hypothesis-generating. The ADHF cohort is single-center, clinical management was not protocolized, and only female pigs were studied (a methodologic constraint of the bladder-cannulation model). Definitive characterization of the dose-response to net fluid removal and of prognostic discrimination at discharge will require an adequately powered, multicenter, prospective study with prespecified thresholds; this is the planned next step.

## Conclusions

Across a clinical cohort and a preload-controlled preclinical model, the locked NIVA Score decreased significantly with acute decongestion, providing direct evidence that the NIVA_HF_ device, already shown to agree with invasively measured PCWP at single time points [3], responds to changes in volume status. These significant, directionally concordant findings support the NIVA Score as a non-invasive marker of congestion and provide the basis for serial monitoring in heart failure, to be confirmed in an adequately powered prospective study.

## Acknowledgments

The authors thank the patients and their families for their participation in this study, the bedside nursing teams who supported the pre- and post-diuresis NIVA acquisitions, and the Vanderbilt animal facility staff who supported the porcine model. The investigators also thank the cardiologists, anesthesiologists, and research coordinators at all participating sites for their collaboration.

## Data availability statement

All data necessary to interpret the findings reported here are presented within the manuscript and its figures. The underlying data cannot be made publicly available because of third-party intellectual-property restrictions: the raw peripheral venous waveforms and the derived NIVA Score outputs constitute protected intellectual property owned by Vanderbilt University Medical Center and licensed to VoluMetrix, LLC, and their public release is prohibited under the governing license and material-transfer agreements. The individual-level clinical waveforms additionally contain potentially identifying patient information and are protected under the governing institutional review board approvals. De-identified data supporting the findings may be made available to qualified investigators for non-commercial research upon reasonable request and execution of a data-use agreement; requests should be directed to the corresponding author. The NIVA algorithm is proprietary and was applied in locked form; inquiries regarding the algorithm should be directed to VoluMetrix, LLC.

## Declaration of generative AI and AI-assisted technologies

During the preparation of this work the authors used a generative AI assistant (Anthropic Claude) to help organize the manuscript and correct grammatical errors. After using this tool, the authors reviewed and edited the content as needed and take full responsibility for the content of the published article. No generative AI was used in the design, performance, analysis, or reporting of the results.

